# Medical concept understanding in large language models is fragmented

**DOI:** 10.64898/2026.03.03.26347552

**Authors:** Lizong Deng, Luming Chen, Mi Liu

**Author notes:** Corresponding authors: Lizong Deng and Luming Chen. These authors contributed equally to this work (Lizong Deng, Luming Chen).

## Abstract

Large language models (LLMs) perform strongly across a wide range of medical applications, yet it remains unclear whether such success reflects genuine understanding of medical concepts. We present an ontology-grounded, concept-centered evaluation of medical concept understanding in LLMs. Using 6,252 phenotype concepts from Human Phenotype Ontology, we decompose concept understanding into three core dimensions—concept identity, concept hierarchy, and concept meaning—and design corresponding benchmarks for each dimension. Across a representative set of contemporary LLMs, best-performing models achieve high accuracy on concept identity (90.6%) and hierarchy (83.8%), but lower performance on concept meaning (72.6%). Concept-level analysis reveals substantial fragmentation in LLM understanding: only 57.7% of concepts are consistently understood across all three dimensions, while 41.3% show partial understanding and 1.1% are not captured in any dimension. These results demonstrate that strong application-level performance of LLMs can mask fundamental gaps in concept-level understanding, highlighting the necessity for ontology-grounded evaluation in medical AI.

## Introduction

Large language models (LLMs) have advanced rapidly in recent years and attracted widespread attention across scientific research and real-world applications^1^. As model capabilities continue to improve, their use in medicine has expanded accordingly, following a clear trajectory from controlled benchmark evaluations toward increasingly realistic clinical settings^2,3^. Representative applications include medical examination benchmarks^4^, clinical question answering systems^5^, and diagnostic assistance tasks^6^. Evaluations on such tasks suggest that LLMs can demonstrate substantial medical competence and, in some scenarios, approach expert-level performance^7^.

However, medical intelligence cannot be fully characterized by success on downstream tasks alone^8,9^. In medicine, knowledge is fundamentally organized around well-defined medical concepts—such as phenotypic abnormalities, disease entities, and therapeutic interventions—which serve as the basic units of clinical observation, diagnosis, and reasoning^10^. For human medical experts, effective clinical reasoning depends not only on producing correct answers in specific situations, but also on a systematic understanding of these concepts and their structured relationships^11^. In this sense, medical intelligence is inherently concept-centered^12^.

Reflecting this conceptual foundation, a large body of medical knowledge has been accumulated, curated, and transmitted in explicitly structured form through biomedical ontologies^13^. Prior to the emergence of LLMs, medical experts formalized foundational medical knowledge primarily by constructing and refining such ontologies. Authoritative resources such as the Unified Medical Language System (UMLS) ^14^ and the Human Phenotype Ontology (HPO) ^15^, developed through decades of sustained expert effort, systematically encode medical concepts together with their synonymous expressions, hierarchical organization, and formal definitions, thereby establishing a stable, explicit, and interpretable medical concept space. These ontologies distill core expert knowledge in medicine and have long provided the basis for concept-based reasoning, knowledge integration, and interpretability in earlier generations of medical artificial intelligence systems^16^.

Against this backdrop, the success of LLMs on downstream medical tasks—such as examinations, clinical question answering, and diagnostic assistance—raises a more fundamental and largely unexplored question: to what extent do LLMs understand basic medical concepts themselves? More specifically, which aspects of medical conceptual knowledge—such as concept identity, concept hierarchy, and concept meaning—are effectively captured by current models, and which remain weakly represented or entirely absent from their internal representations (**Fig. 1**)? This question is particularly consequential because, for human experts, medical concepts form the foundation of clinical reasoning. If LLMs achieve strong task-level performance without forming robust and systematic concept-level understanding, the reliability, interpretability, and generalizability of their downstream behavior warrant careful scrutiny^17^.

**Figure 1.**
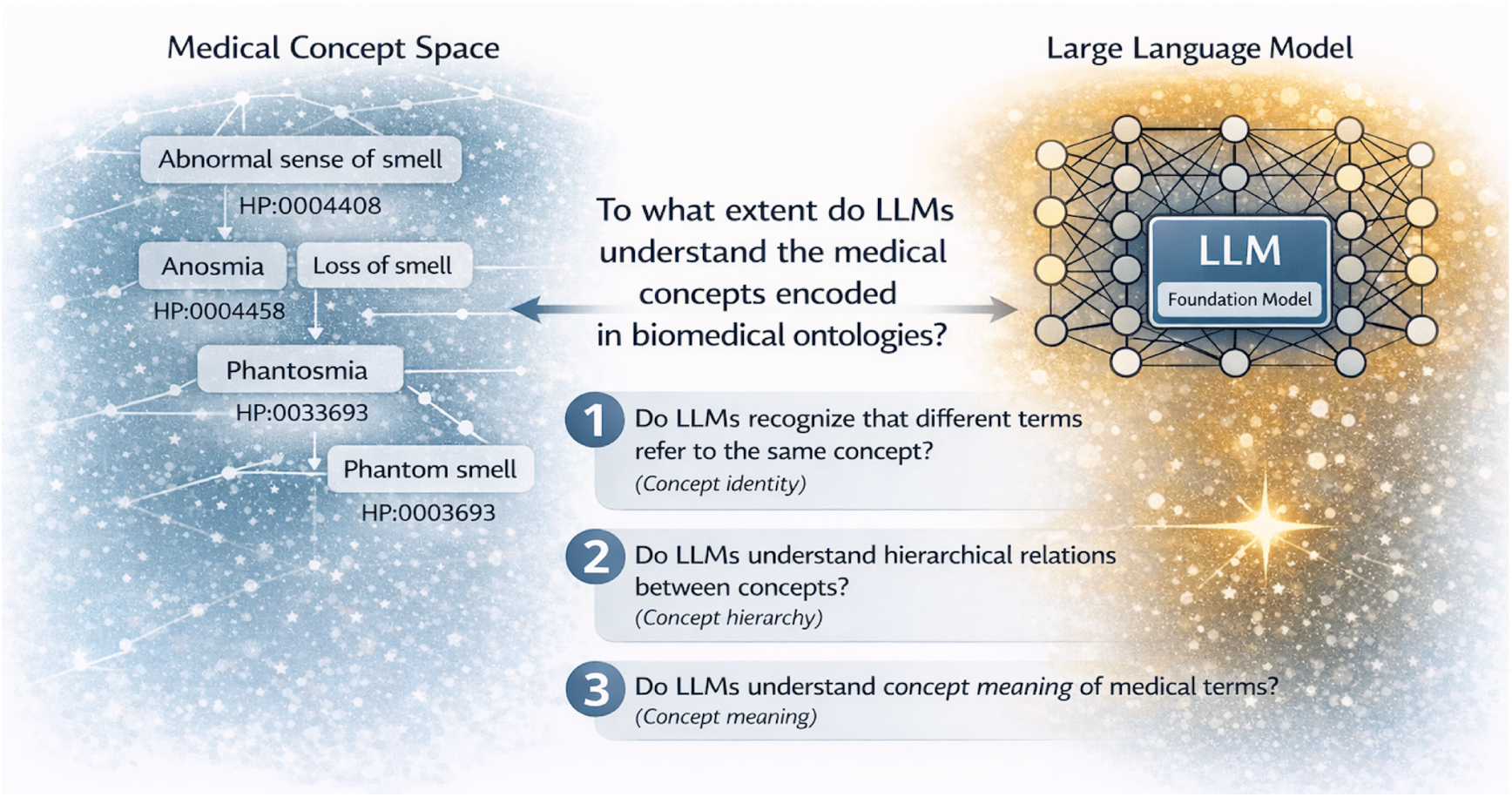
Ontology-grounded framing of medical concept understanding in large language models.

Despite the central importance of this issue for the future of medical artificial intelligence, existing research has not yet provided a systematic answer. Prior work has largely examined the conceptual capabilities of LLMs indirectly through task-oriented evaluations. One line of research investigates whether LLMs can recognize or extract medical concepts from unstructured clinical or biomedical text, such as identifying phenotype mentions in clinical narratives or biomedical abstracts^18–20^. These studies demonstrate that LLMs can capture phenotype-related expressions and extract medically relevant information from text, but their evaluations rely primarily on extraction performance and offer limited insight into models’ understanding of underlying conceptual structure. A second line of research focuses on medical entity linking, assessing whether surface-level medical terms can be correctly mapped to standardized entries in biomedical ontologies such as the Human Phenotype Ontology^21–23^. While these studies examine alignment between linguistic expressions and ontological identifiers, their primary emphasis remains on task-level accuracy rather than on whether models form systematic concept-level understanding of different types of ontological knowledge, including synonym structure, hierarchical relations, and formal definitions.

Overall, existing work provides strong evidence that LLMs can operate over medical concepts in concrete downstream tasks, but it does not directly address a more fundamental question: to what extent LLMs truly understand the medical concepts encoded in biomedical ontologies, and how consistently such understanding is distributed across different conceptual dimensions. Current evaluations are typically task-specific, rely on relatively limited datasets, and do not systematically cover the full scope of conceptual knowledge accumulated in authoritative biomedical ontologies. In particular, few studies assess models simultaneously across concept identity, hierarchical structure, and conceptual meaning. As a result, it remains unclear whether LLMs possess coherent and integrated medical concept understanding, or whether strong application-level performance masks substantial fragmentation at the conceptual level.

In this work, we address this gap through an ontology-grounded, concept-centered evaluation framework. We decompose medical concept understanding into three foundational dimensions—concept identity, concept hierarchy, and concept meaning—which together reflect the core organizational principles of biomedical ontologies. Based on these dimensions, we construct three complementary concept-level evaluation tasks to assess synonym recognition, hierarchical reasoning, and definition-level understanding. Using the Human Phenotype Ontology as a controlled and authoritative conceptual framework, we curate benchmark datasets grounded in explicitly defined medical concepts and relations, and systematically evaluate a diverse set of state-of-the-art commercial models, open-source models, and domain-specialized medical LLMs. Through this analysis, we move beyond application-level performance and provide a principled assessment of the extent to which current large language models understand medical concepts.

## Results

To systematically examine the extent to which LLMs understand medical concepts, we conducted three ontology-grounded computational experiments that probe progressively deeper levels of conceptual understanding. Specifically, these experiments assess concept identity through synonym recognition, concept hierarchy through hypernym recognition, and concept meaning through definition sentence selection. We evaluated a representative set of contemporary LLMs spanning three major categories, including state-of-the-art commercial models (Gemini-3 Flash and GPT-5.2), state-of-the-art open-source models (Qwen3-235B and DeepSeek-3.2), and domain-knowledge–tuned medical language models (Baichuan-M2 and Citrus-Qwen72B). Both Baichuan-M2 and Citrus-Qwen72B are derived from the same base model, Qwen2.5-72B, through fine-tuning on large-scale medical corpora. To enable direct comparison between domain-tuned models and their underlying foundation model, Qwen2.5-72B was also included in all experiments. All experiments are constructed over the same set of medical concepts drawn from the Human Phenotype Ontology. The detailed results of these experiments are reported below.

### Concept identity is robustly captured but unevenly distributed across LLMs

The first experiment examined whether LLMs can recognize that different medical terms refer to the same underlying medical concept, corresponding to concept identity in biomedical ontologies. For example, the terms “*anosmia*” and “*loss of smell*” are treated as synonyms in the Human Phenotype Ontology and are both mapped to the same concept identifier (HP:0000458). To probe this capability, models were evaluated on a controlled synonym recognition task constructed from the Human Phenotype Ontology, comprising 6,252 phenotype concepts. Each query required selecting the correct synonym from three carefully designed distractors, including a sibling concept within the ontology, a morphologically similar but non-synonymous term, and a randomly sampled medical concept (*Fig. 2a*, see *Methods* for details).

**Figure 2.**
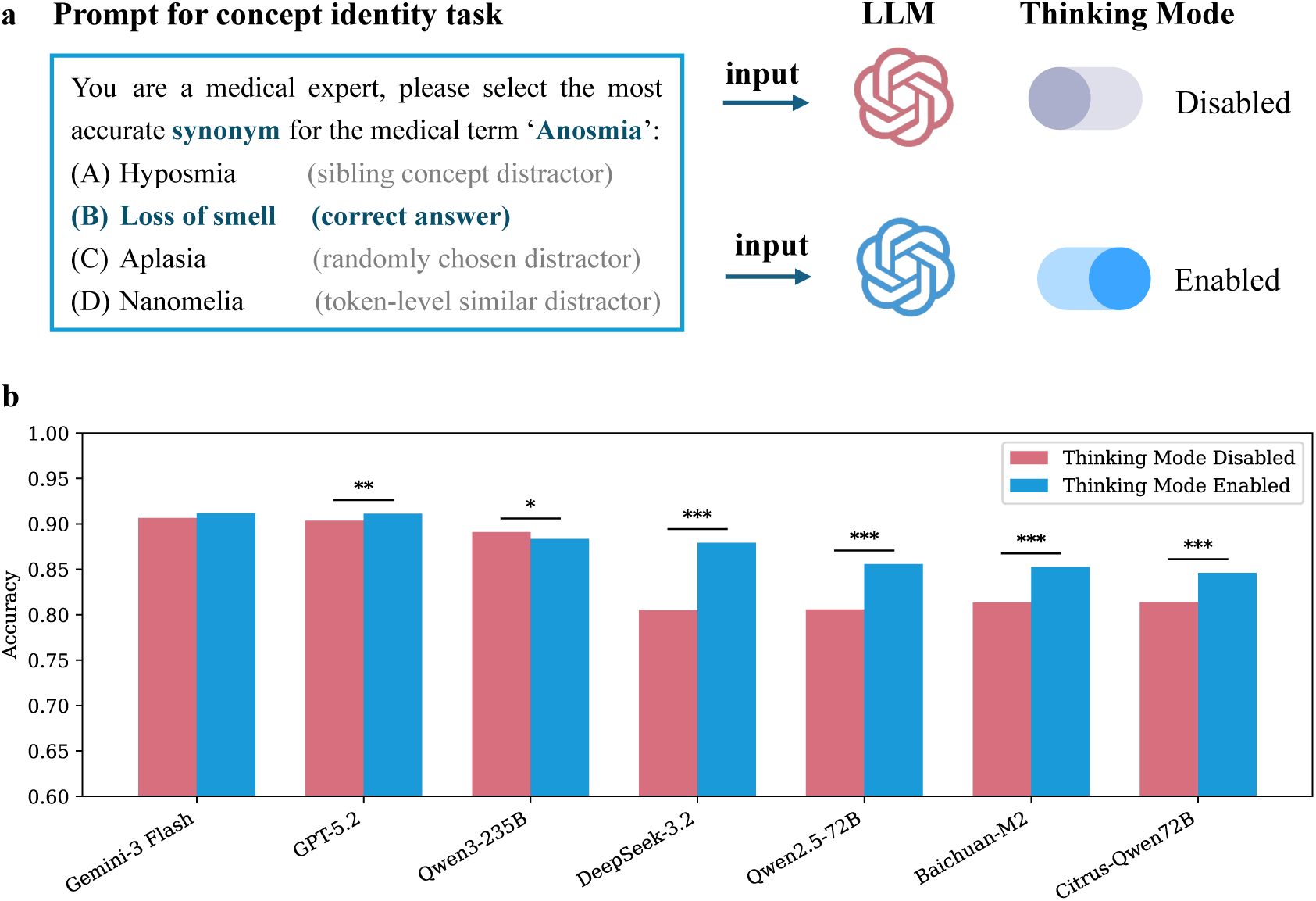
Concept identity understanding in LLMs under standard and reasoning-enabled inference. (a) Example prompt for the concept identity task. Given a medical term (e.g., Anosmia), models were required to select the most accurate synonym from four candidate options. Example prompt for the concept identity task. Given a medical term (e.g., Anosmia), models were required to select the most accurate synonym from four candidate options. Each query was evaluated under two inference conditions: standard inference (thinking mode disabled) and reasoning-enabled inference (thinking mode enabled). (b) Accuracy of seven LLMs on the concept identity task with thinking mode disabled (red) and enabled (blue). Bars indicate mean accuracy across all evaluated phenotype concepts. Statistical significance of performance differences between the two settings was assessed using McNemar’s test with Bonferroni correction (* *p* < 0.05, ** *p* < 0.01, *** *p* < 0.001).

All evaluated models support an optional thinking mode, a reasoning-enabled setting that encourages more explicit, step-by-step internal deliberation during inference, trading additional computation time for potentially more accurate and reliable responses on complex tasks. Because such explicit reasoning could, in principle, improve the recognition of subtle synonym relations, we evaluated model performance under both standard settings (thinking mode disabled) and reasoning-enabled settings (thinking mode enabled) (*Fig. 2a*).

*Fig. 2b* summarizes the performance of the seven evaluated LLMs on the synonym recognition task, using accuracy as the evaluation metric. Each model was evaluated under both standard settings (thinking mode disabled) and reasoning-enabled settings (thinking mode enabled). Across models, performance on the synonym recognition task was consistently high, indicating that synonym-level concept identity is generally well captured by contemporary LLMs.

Under standard settings without thinking mode, the top-performing models were the commercial general-purpose LLMs Gemini-3 Flash and GPT-5.2, together with the open-source model Qwen3-235B, achieving accuracies of 90.6%, 90.3%, and 89.1%, respectively. In contrast, the two domain-knowledge–tuned medical models, Baichuan-M2 and Citrus-Qwen72B, achieved accuracies of approximately 81.4%. Although this represents a modest improvement over their shared base model Qwen2.5-72B (80.6%), their performance remained substantially below that of leading general-purpose models such as Gemini-3 Flash (p < 0.001, McNemar’s test).

Enabling thinking mode generally led to performance improvements, but the magnitude and direction of this effect varied markedly across models (*Fig. 2b*). For Gemini-3 Flash, accuracy increased modestly from 90.6% to 91.2%. In contrast, Qwen3-235B exhibited a slight decrease in performance when thinking mode was enabled, with accuracy dropping from 89.1% to 88.4%. For DeepSeek-3.2, however, thinking mode resulted in a pronounced improvement, with accuracy increasing from 80.5% to 87.9%. The two domain-knowledge–tuned medical models also benefited from thinking mode, with Baichuan-M2 improving from 81.4% to 85.2% and Citrus-Qwen72B from 81.3% to 84.6%. Nevertheless, even under reasoning-enabled settings, their performance did not surpass that of the strongest general-purpose LLMs.

Overall, these results indicate that synonym-level concept identity is robustly captured by LLMs, but that performance is unevenly distributed across model families and influenced in a model-dependent manner by reasoning-enabled inference.

### Concept hierarchy understanding is markedly weaker than concept identity in LLMs

Having established that LLMs can reliably recognize when different medical terms refer to the same underlying concept, we next examined whether this capability extends to understanding how medical concepts are organized within an ontological hierarchy. This experiment probes concept hierarchy understanding, which requires models to identify is-a relations that place a concept within a structured taxonomy, rather than merely associating alternative lexical expressions. For example, in the Human Phenotype Ontology, the concept “*Anosmia*” (HP:0000458) is defined as a child of the more general concept “*Abnormality of the sense of smell*” (HP:0004408).

Models were evaluated on a controlled hypernym recognition task derived from the Human Phenotype Ontology, using the same set of medical concepts as in the synonym recognition experiment. For each query term, models were asked to select the most appropriate parent concept from three carefully constructed distractors, including a sibling concept within the ontology, a morphologically similar but ontologically unrelated concept, and a randomly sampled concept (*Fig. 3a*; see Methods for details). As in the synonym recognition task, all models were evaluated under both standard settings and reasoning-enabled settings (thinking mode).

**Figure 3.**
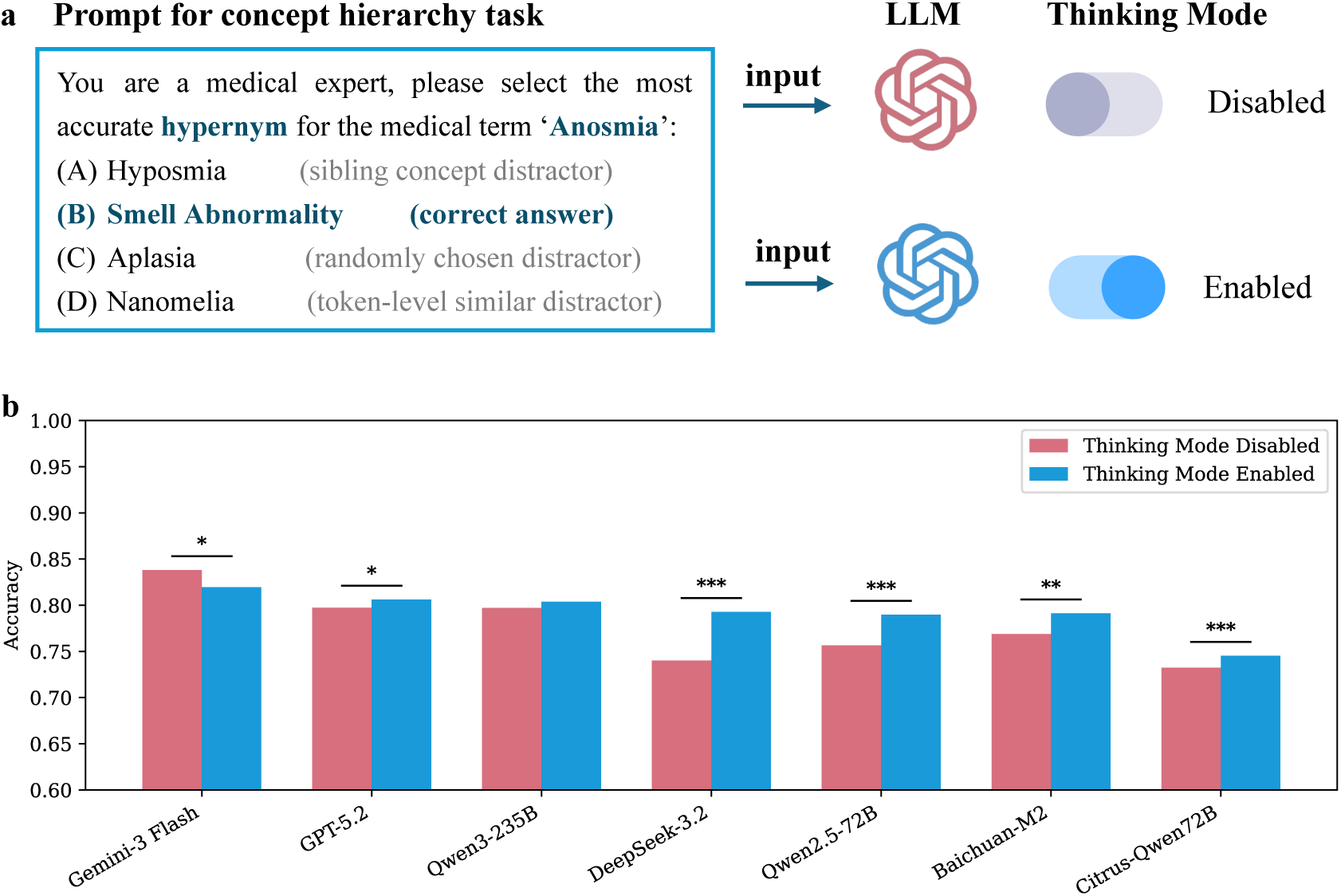
Concept hierarchy understanding in LLMs under standard and reasoning-enabled inference. (a) Example prompt for the concept hierarchy task. Given a medical term (e.g., Anosmia), models were required to select the most accurate hypernym from four candidate options. Each query was evaluated under two inference conditions: standard inference (thinking mode disabled) and reasoning-enabled inference (thinking mode enabled). (b) Accuracy of seven LLMs on the concept hierarchy task under standard (red) and reasoning-enabled (blue) inference. Bars indicate mean accuracy across all evaluated phenotype concepts. Statistical significance of performance differences between inference conditions was assessed using McNemar’s test with Bonferroni correction (* *p* < 0.05, ** *p* < 0.01, *** *p* < 0.001).

*Fig. 3b* summarizes model performance on the concept hierarchy understanding task, with accuracy as the evaluation metric. Under standard settings without thinking mode, the top-performing models remained Gemini-3 Flash, GPT-5.2, and Qwen3-235B, achieving accuracies of 83.8%, 79.7%, and 79.7%, respectively. The two domain-knowledge–tuned medical models, Baichuan-M2 and Citrus-Qwen72B, achieved accuracies of 76.8% and 73.2%, respectively, while their shared base model Qwen2.5-72B achieved an accuracy of 75.6%. The impact of thinking mode on hierarchical understanding was heterogeneous across models (*Fig. 3b*). For Gemini-3 Flash, enabling thinking mode led to a modest but statistically significant decrease in accuracy, from 83.8% to 82.0% (*p* < 0.05, McNemar’s test). In contrast, Qwen3-235B showed no significant change in performance, with accuracy remaining stable (79.7% vs. 80.3%); while DeepSeek-3.2 exhibited a larger gain, improving from 74.0% to 79.2% (*p* < 0.001, McNemar’s test). Similar moderate improvements were observed for the domain-knowledge–tuned models, with Baichuan-M2 increasing from 76.8% to 79.1% and Citrus-Qwen72B from 73.2% to 74.5%. Despite these gains, domain-specialized medical models continued to underperform relative to leading general-purpose LLMs on this task (*p* < 0.001, McNemar’s test).

Although these results indicate that contemporary LLMs possess a non-trivial capacity to reason about hierarchical relations, performance was consistently lower than that observed for synonym-level concept identity (*Fig. 4*). For instance, without thinking mode enabled, Gemini-3 Flash exhibited a decrease of 6.8 percentage points relative to its synonym recognition accuracy (90.6%), whereas GPT-5.2 showed a larger decline of 10.6 percentage points (*p* < 0.001 for both comparisons, McNemar’s test).

**Figure 4.**
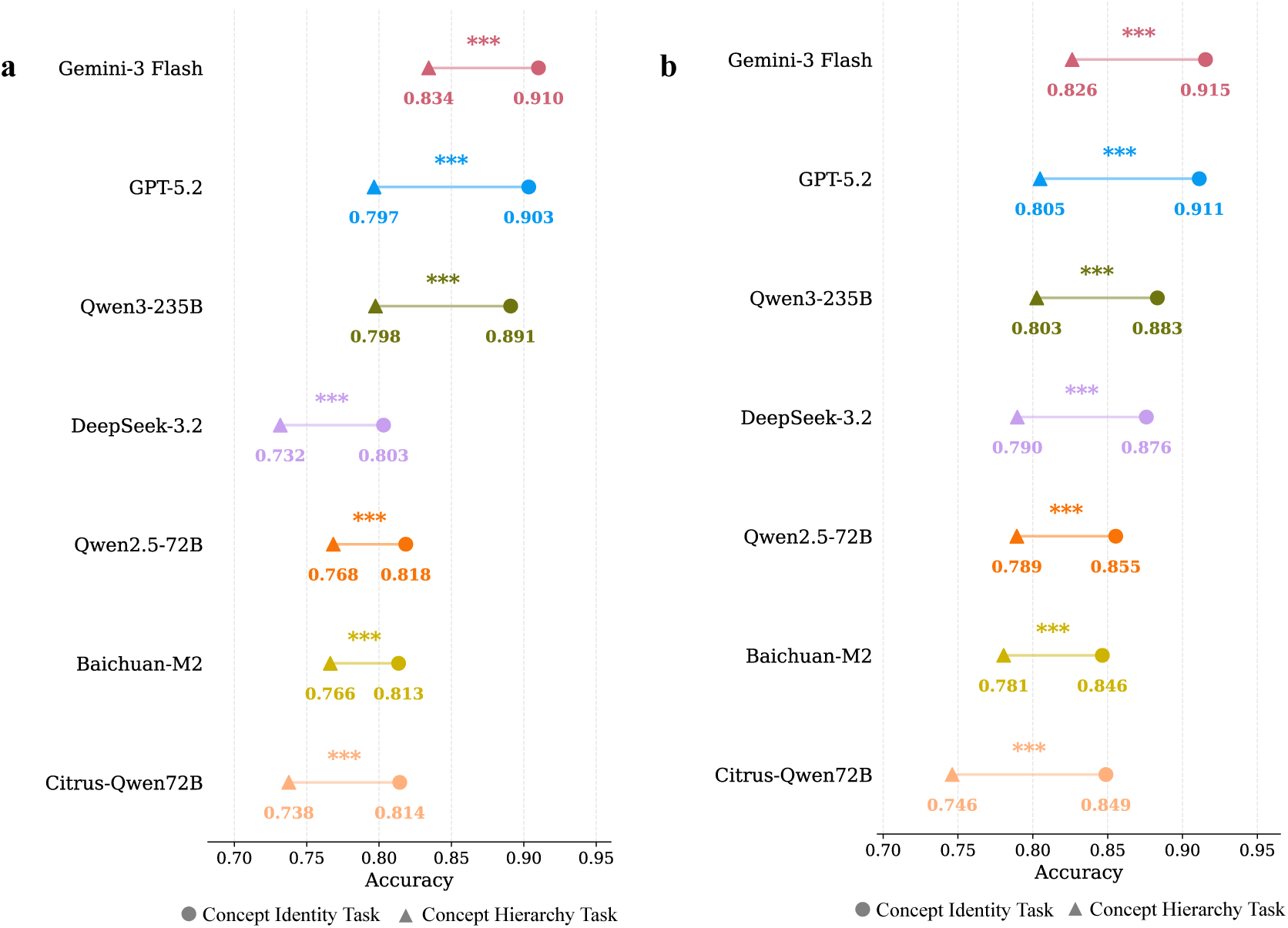
Task-level comparison between concept identity and concept hierarchy understanding in LLMs. (a) Model accuracy on synonym recognition (concept identity) and hypernym selection (concept hierarchy) with thinking mode disabled. (b) Model accuracy on synonym recognition (concept identity) and hypernym selection (concept hierarchy) with thinking mode enabled. Statistical significance was assessed using McNemar’s test with Bonferroni correction (\**p* < 0.05, ** *p* < 0.01, *** *p* < 0.001).

Overall, although LLMs demonstrate reasonably strong performance on concept hierarchy understanding—reaching a maximum accuracy of 83.8%—their performance consistently declines relative to synonym-level concept identity recognition. This systematic reduction, observed across all evaluated model families, highlights a clear and reproducible gap between recognizing concept identity and correctly situating concepts within an ontological hierarchy.

### Concept meaning understanding is highly sensitive to contextual knowledge in LLMs

We next examined whether LLMs understand what a medical concept means, beyond recognizing alternative names or its position within an ontological hierarchy. This experiment probes definition-level concept meaning, requiring models to identify the correct semantic description of a medical concept from a set of candidate definitions. For example, given a query such as “*Loss of smell*” (a non-preferred synonym of *Anosmia*), models were asked to select, from twenty candidate definition sentences, the definition that best captures the meaning of the underlying concept (*Fig. 5a*). All candidate definitions were presented in a standardized form (“The definition of [the concept’s preferred term] is …”); in this case, the correct definition is expressed using the preferred term “*Anosmia*”, rather than the query term “*Loss of smell*” itself. Query terms were deliberately drawn from non-preferred synonyms to increase semantic difficulty and prevent trivial surface-form matching between the query and candidate definitions.

**Figure 5.**
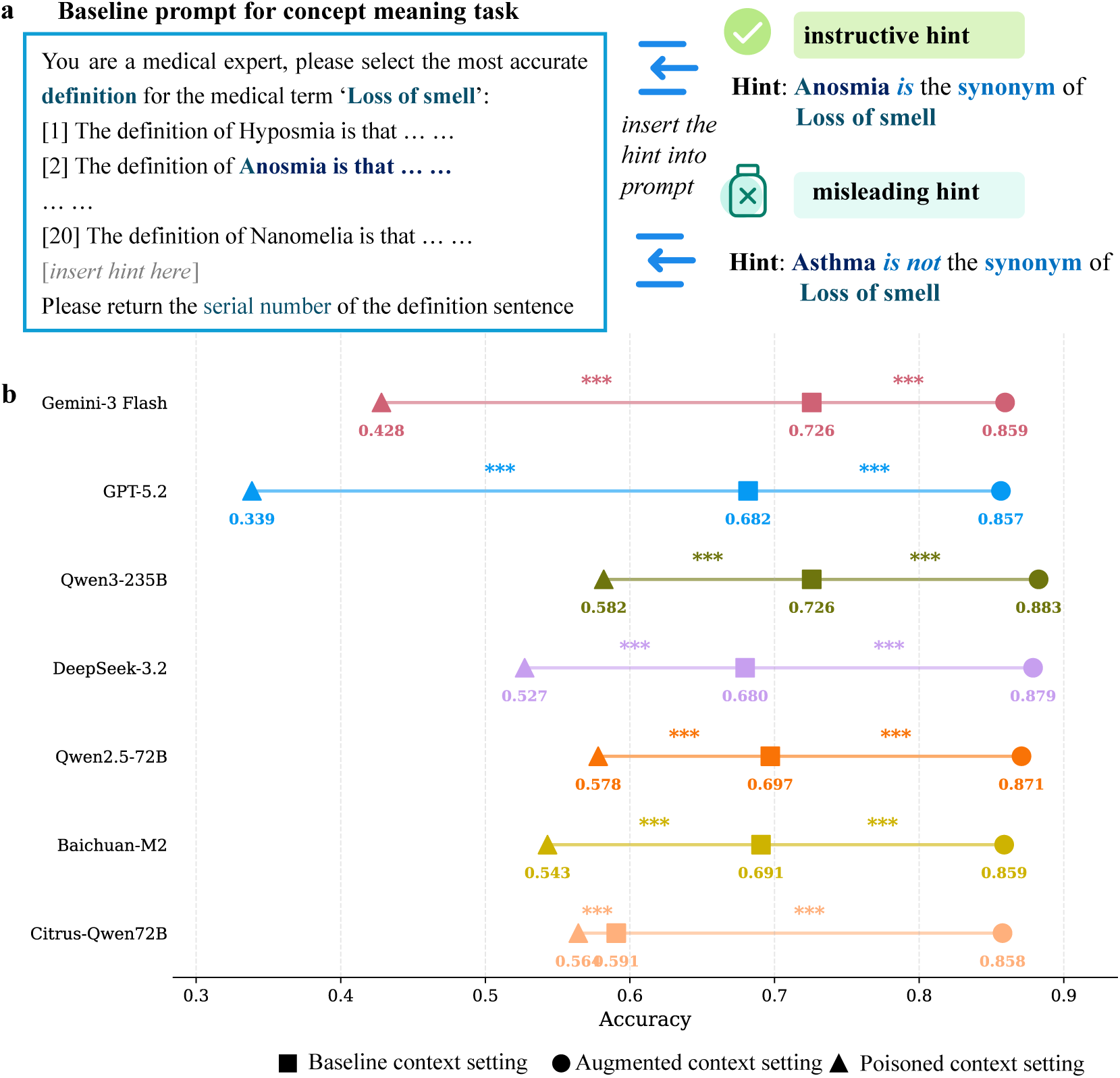
Concept meaning understanding in LLMs under different contextual settings. (a) Example prompt for the concept meaning task. Given a medical query term drawn from a non-preferred synonym (e.g., Loss of smell), models were required to select the correct definition of the underlying medical concept from twenty candidate definitions. Each query was evaluated under three context conditions: a baseline setting with no additional information, an augmented setting providing correct synonym guidance, and a poisoned setting providing incorrect synonym guidance. (b) Accuracy of seven LLMs on the concept meaning task under baseline (square), augmented (circle), and poisoned (triangle) context settings. Points indicate mean accuracy across all evaluated phenotype concepts. Statistical significance of performance differences between context conditions was assessed using McNemar’s test with Bonferroni correction (* *p* < 0.05, ** *p* < 0.01, *** *p* < 0.001).

Under this task formulation, model performance was evaluated under three parallel context conditions (*Fig. 5a*). In the baseline context setting, no explicit ontological guidance was provided. In the augmented context setting, the prompt explicitly indicated that the query term (e.g., *Loss of smell)* and the preferred term appearing in the definition sentence (e.g., *Anosmia*) refer to the same medical concept. In contrast, in the poisoned context setting, the prompt explicitly stated that the query term and the preferred term in the definition sentence *do not* have a synonymous relationship. This design enables a direct comparison of model behavior under neutral, supportive, and misleading contextual knowledge.

*Fig. 5b* summarizes model performance on the concept meaning understanding task across the three context conditions. Under the baseline context setting, overall performance was moderate and exhibited notable variation across models. The top-performing systems were Gemini-3 Flash (72.6%), Qwen3-235B (72.5%), and Qwen2.5-72B (69.7%). GPT-5.2, which ranked near the top in the previous two tasks, dropped to fifth place with an accuracy of 68.2%. The domain-knowledge–tuned model Baichuan-M2 achieved a comparable performance (69.1%), whereas Citrus-Qwen72B performed substantially worse, with an accuracy of 59.1%. This shift in model ranking relative to the synonym and hierarchy tasks indicates that definition-level concept understanding reflects a distinct and more variable capability profile.

When accurate ontological guidance was provided in the augmented context setting, performance improved substantially across all evaluated models (*Fig. 5b*). Under this condition, Qwen3-235B achieved the highest accuracy (88.2%), followed closely by DeepSeek-3.2 (87.8%) and Qwen2.5-72B (87.1%). Gemini-3 Flash, GPT-5.2, Baichuan-M2, and Citrus-Qwen72B exhibited similar performance levels, with accuracies ranging from 85.7% to 85.9%. These results demonstrate that accurate external ontological information can strongly enhance definition-level performance across diverse model architectures. Notably, however, even under this favorable condition, performance remained well below perfect accuracy, indicating that models did not fully exploit the provided guidance to achieve stable concept meaning understanding.

In contrast, when incorrect ontological information was supplied in the poisoned context setting, performance deteriorated sharply across all models (*Fig. 5b*). Under this condition, the highest accuracy was achieved by Qwen3-235B (58.2%), while GPT-5.2 ranked last with an accuracy of 33.9%. Similar reductions were observed for other models, collectively demonstrating that definition-level concept understanding is highly sensitive to the correctness of contextual knowledge. Importantly, despite the presence of misleading guidance, performance did not collapse to chance level, suggesting that LLMs were not uniformly misled by incorrect external information.

Overall, under the baseline setting, LLMs demonstrated a meaningful—though limited—ability to identify definition-level descriptions of medical concepts (e.g., 72.6% accuracy for Gemini-3 Flash). Besides, performance on this task was highly sensitive to contextual knowledge: accurate ontological guidance substantially improved accuracy without resolving the task, whereas misleading guidance caused pronounced degradation. Together, these findings indicate that definition-level medical concept understanding in LLMs remains fragile and heavily dependent on external contextual support rather than stable, internally grounded semantic representations.

### Concept-level analysis reveals fragmented concept understanding in LLMs across concept identity, hierarchy, and meaning

To integrate the findings from the three preceding experiments, we conducted a concept-level analysis to assess how consistently individual medical concepts are understood across the three dimensions of concept identity, concept hierarchy, and concept meaning. This analysis focused on Gemini-3 Flash, which achieved the strongest overall performance across all three tasks. For each of the 6,252 medical concepts evaluated, we examined whether the model correctly answered the corresponding synonym recognition, hierarchy recognition, and definition understanding questions. Based on this per-concept evaluation, concepts were categorized into three groups: fully understood concepts, for which all three dimensions were answered correctly; partially understood concepts, for which one or two dimensions were answered correctly; and poorly understood concepts, for which the model failed on all three dimensions.

This analysis revealed that 57.7% of concepts were consistently understood across all three dimensions (*Fig. 6a*). By contrast, 41.3% of concepts exhibited only partial understanding, indicating fragmented representations spanning concept identity, hierarchy, and meaning. Notably, only 1.1% of concepts were not understood in any dimension by Gemini-3 Flash.

**Figure 6.**
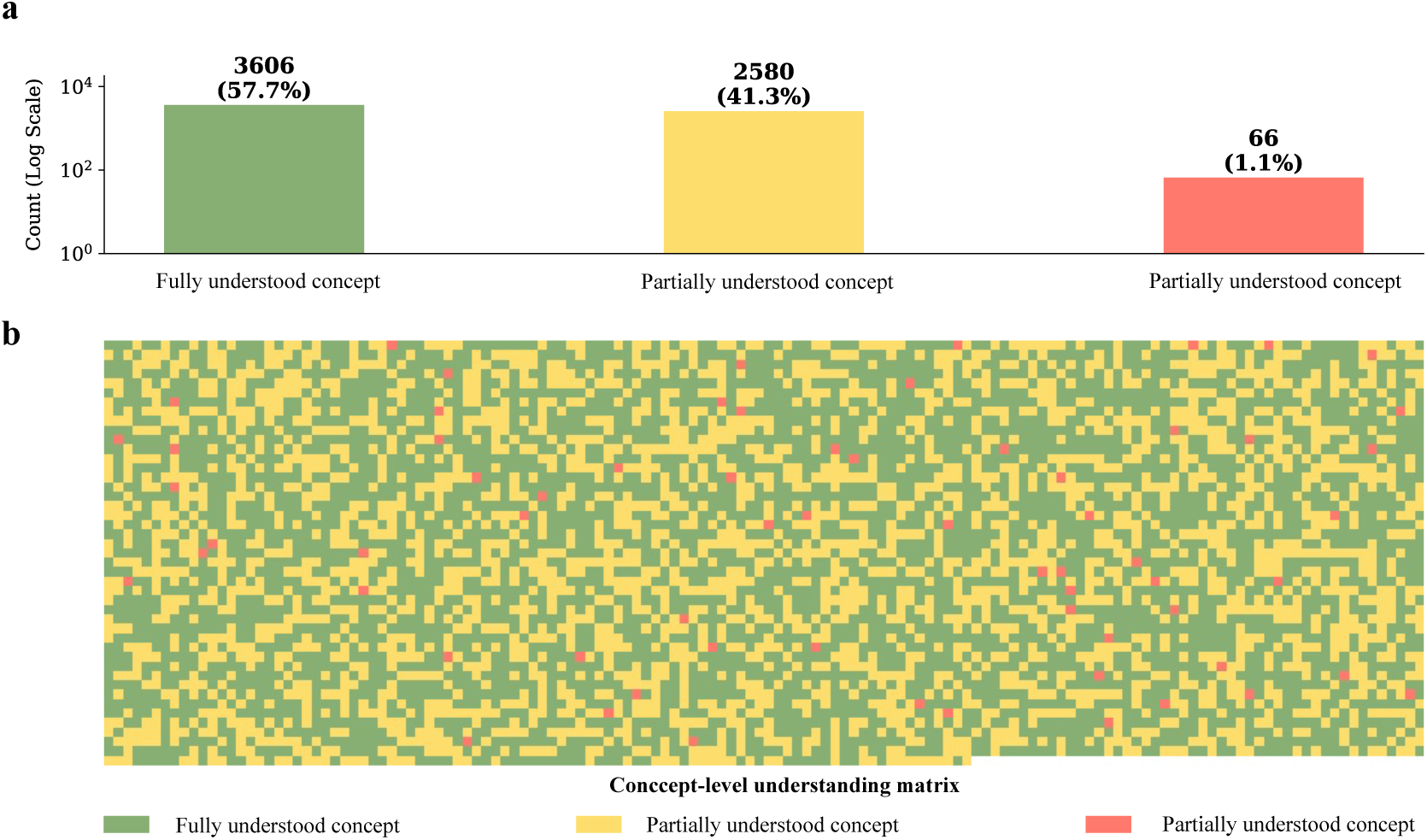
Fragmented medical concept understanding at the concept level in LLMs. (a) Concept-level distribution of medical concept understanding for the best-performing model, Gemini-3 Flash. Each of the 6,252 phenotype concepts was evaluated across three dimensions—concept identity, concept hierarchy, and concept meaning—and classified as fully understood (all three correct), partially understood (one or two correct), or poorly understood (all three incorrect). Counts and proportions are shown on a logarithmic scale. (b) Concept-level understanding matrix across the full medical concept space. Each cell represents a single medical concept, colored by understanding status (green, fully understood; yellow, partially understood; red, poorly understood).

To visualize this pattern, we constructed a concept-level map of model understanding that highlights fragmentation across dimensions (*Fig. 6b*). The full set of 6,252 concepts was arranged in a matrix, with each cell representing a single medical concept. Fully understood concepts are shown in green, partially understood concepts in yellow, and poorly understood concepts in red. Rather than forming coherent or contiguous regions, these categories are interspersed throughout the matrix, revealing a highly heterogeneous and fragmented distribution of concept understanding across the medical concept space. This visualization provides an intuitive, concept-level depiction of fragmented medical concept understanding in LLMs.

## Discussions

The present study provides a systematic, ontology-grounded examination of medical concept understanding in LLMs by decomposing this capacity into three foundational dimensions—concept identity, concept hierarchy, and concept meaning. Across a diverse set of contemporary LLMs, we observe a consistent pattern: models that achieve strong performance on a wide range of medical applications do not exhibit uniformly strong understanding across these conceptual dimensions. Instead, medical concept understanding is unevenly distributed, both across dimensions and across individual concepts, resulting in fragmented conceptual profiles rather than coherent, integrated representations.

At a mechanistic level, the fragmented pattern observed here can be plausibly explained by how LLMs are trained, namely through next-token prediction^24^. This training objective rewards models for producing locally plausible continuations of text, rather than for constructing globally consistent conceptual representations^17^. As a result, LLMs tend to learn patterns that are repeatedly and explicitly reinforced in language. In medical text, synonymy is often expressed through paraphrase and co-occurrence, making identity-level associations relatively easy to acquire^25^. By contrast, hierarchical relations and formal definitions rely on structured constraints that are rarely stated explicitly and consistently in natural language, providing a much weaker learning signal. Consequently, models can perform well on many medical tasks while still lacking a coherent, ontology-aligned representation of the underlying concept space^2,26^.

Viewed from another perspective, these results also underscore the non-trivial conceptual capacities that can emerge from language-only training^24^. In our analysis of Gemini-3 Flash, 57.7% of medical concepts were consistently captured across all three dimensions, with a further 41.3% exhibiting partial understanding, despite the absence of any explicit ontological supervision. This degree of concept acquisition highlights the remarkable ability of LLMs to distill structured conceptual knowledge from unstructured text, reflecting the emergence of broadly applicable general intelligence^27,28^. At the same time, medicine is a domain in which conceptual precision is not merely advantageous but essential. Fragmented understanding of foundational medical concepts may compromise the rigor and faithfulness of clinical reasoning, with direct implications for the reliability and safety of downstream clinical deployment^29,30^.

This limitation becomes particularly apparent in the strong sensitivity of definition-level concept understanding to external context. When accurate ontological cues are provided, model performance improves markedly; when misleading cues are introduced, performance declines sharply. This contrast indicates that definition-level meaning is not robustly encoded as a stable internal representation, but instead depends heavily on contextual support^31,32^. Notably, performance does not fall to chance even under incorrect guidance, suggesting that models retain partial semantic constraints rather than being fully driven by context. Together, these findings point to a hybrid representational regime in which data-driven learning enables flexible semantic associations, yet fails to produce the stable, internally coherent conceptual structure characteristic of explicitly organized knowledge. This pattern is consistent with recent evidence showing that medical LLMs can be highly sensitive to subtle perturbations in training data or contextual prompts, including data poisoning and prompt injection^33,34^.

These findings have important implications for medical artificial intelligence. Strong performance on application-level benchmarks can mask systematic limitations in concept-level understanding, with consequences for robustness, interpretability, and generalization. Rather than diminishing the role of ontologies, the apparent success of LLMs may in fact heighten the need for explicit conceptual grounding as a stabilizing substrate for medical reasoning. From this perspective, integrating language models with ontology-based representations may be critical for developing medical AI systems that are not only high-performing, but also conceptually coherent and trustworthy^2,35–37^.

Several limitations of this study should be acknowledged. Although biomedical ontologies cover a wide range of medical knowledge, our analysis focuses on phenotype concepts from the Human Phenotype Ontology. This choice was motivated by both conceptual and practical considerations: phenotypic abnormalities are central to clinical observation and diagnosis, and their scale allows controlled, ontology-grounded evaluation at manageable computational cost. Nonetheless, other medical domains—such as diseases, procedures, or pharmacological entities—may exhibit similar or different patterns of concept understanding in LLMs. Extending the present framework to these domains remains a promising avenue for future investigation.

## Methods

### Ontology selection and conceptual grounding

To ground our evaluation of medical concept understanding in a principled and well-defined conceptual framework, we selected the Human Phenotype Ontology (HPO) as the ontological resource underlying all experiments. HPO is a curated biomedical ontology that systematically represents abnormal human phenotypes, including standardized definitions, synonymous expressions, and hierarchical relations, making it well suited for controlled analyses of medical concept structure^15^.

In principle, large-scale biomedical ontologies such as the Unified Medical Language System (UMLS) ^14^ could be used to study medical concept understanding. Indeed, UMLS was initially considered due to its broad coverage of biomedical knowledge. However, the primary consideration guiding our final choice was experimental feasibility at scale. At the time of this study, UMLS (release 2025AA) comprised more than 3.45 million concepts spanning highly heterogeneous biomedical vocabularies. Conducting controlled, ontology-grounded evaluations over such a large and diverse concept space would require substantially greater computational resources and incur prohibitive computational and associated costs, making systematic concept-level experimentation impractical within the scope of this work.

Given these constraints, we turned to HPO as a focused yet conceptually rich alternative. Phenotypic abnormalities constitute one of the most fundamental categories of medical concepts, forming the basis of clinical observation, diagnosis, and therapeutic decision-making across medical domains. By concentrating on phenotypes, our evaluation targets a core layer of medical conceptual knowledge while retaining sufficient structural complexity to probe concept identity, hierarchy, and meaning in a controlled manner. Moreover, HPO offers a favorable balance between conceptual expressiveness and tractability: the HPO 2025-09-01 release contains 11,547 phenotype concepts, enabling systematic experimentation without sacrificing interpretability.

Taken together, these considerations make HPO a methodologically appropriate and practically viable choice for investigating how large language models represent medical concepts across multiple conceptual dimensions.

### Concept selection and concept set construction

Starting from the full HPO release comprising 11,547 phenotype concepts, we curated a concept set to support controlled evaluation of medical concept understanding across identity, hierarchy, and meaning. This process focused on selecting concepts that (i) possess non-trivial synonymous variation, (ii) minimize reliance on superficial lexical similarity, and (iii) remain suitable for consistent evaluation across multiple conceptual dimensions. The selection procedure consisted of three sequential steps.

1. Concept eligibility and synonym-based screening. Among all HPO concepts, 11,309 were associated with at least one recorded synonym in addition to a preferred term. Only these concepts were considered further, as synonym information is a prerequisite for evaluating concept identity. For each eligible concept, we computed the lexical similarity between its preferred term and each associated synonym using the Token Set Ratio metric implemented in the *thefuzzy* Python package. To conservatively characterize synonym variability, we retained, for each concept, the synonym with the *lowest* Token Set Ratio relative to the preferred term, corresponding to the most lexically dissimilar synonym.
2. Lexical similarity filtering. To reduce the likelihood that models could succeed through trivial string matching or surface-level lexical overlap, we excluded concepts whose least-similar synonym remained highly similar to the preferred term. Specifically, concepts were filtered using a threshold of Token Set Ratio < 80, removing cases in which synonyms differed only marginally (e.g., pluralization or word order variation). After this filtering step, 6,656 concepts remained, representing cases in which synonym recognition requires abstraction beyond simple lexical similarity.
3. Concept feasibility filtering across ontological structure. From the filtered concept pool, we further excluded concepts that could not support consistent evaluation across all conceptual dimensions due to structural limitations in the ontology (e.g., missing suitable parent or sibling concepts). This feasibility filtering ensured that each retained concept could be uniformly evaluated in subsequent experiments on concept identity, hierarchy, and meaning. After this step, 6,252 medical concepts were retained and used consistently across all experiments reported in this study.

### Task formulation for concept identity understanding

The concept identity understanding task was designed to assess whether LLMs can recognize that different medical terms refer to the same underlying medical concept, independent of surface-level lexical similarity. This task operationalizes concept identity as defined in biomedical ontologies, in which multiple synonymous expressions are mapped to a single concept identifier.

Building on the curated concept set described above, we formulated this task as a controlled multiple-choice synonym recognition problem grounded in the Human Phenotype Ontology. For each selected concept, a synonym recognition instance was constructed using a standardized prompt of the form:

> “*You are a medical expert. Please select the most accurate synonym for the medical term ‘[query term]’*.”

Each instance consisted of four candidate answer options, exactly one of which corresponded to a true synonym of the underlying concept. The construction of both correct answers and distractors followed a set of explicit design principles, described below.

1. Selection of lexically distinct true synonyms. To reduce the likelihood that models could rely on trivial string matching or surface-level lexical overlap, the correct answer for each instance was deliberately chosen to be a lexically distant synonym of the concept’s preferred term. Specifically, for each concept, we identified the synonym exhibiting the lowest token-based lexical similarity to the preferred term, subject to the similarity threshold described above. This design aims to minimize the extent to which correct answers can be recovered through superficial lexical matching.
2. Distractor design. Each instance included three distractors designed to probe distinct failure modes in concept identity recognition: (i) a sibling distractor, drawn from a concept that shares the same parent in the HPO hierarchy, introducing semantic proximity without identity equivalence; (ii) a lexical distractor, lexically similar to the preferred term but referring to a different medical concept, designed to mislead models relying on surface-form similarity; and (iii) a random distractor, sampled from an unrelated concept, serving as a baseline control. Together, these distractors create an adversarial setting in which lexically similar but semantically incorrect options compete directly with lexically dissimilar but semantically correct answers.
3. Task format. All instances were formatted as four-option multiple-choice questions using a fixed and standardized prompt template. The order of answer options was randomized for each instance. Models were instructed to return the option corresponding to the most accurate synonym.

### Task formulation for concept hierarchy understanding

The concept hierarchy understanding task was designed to assess whether large language models can correctly recognize ontological is-a relationships between medical concepts. Unlike concept identity, which concerns whether different terms refer to the same concept, this task probes whether models can situate a concept within a structured conceptual taxonomy and identify its appropriate parent concept.

Building on the curated concept set described above, we formulated this task as a controlled multiple-choice hypernym recognition problem grounded in the hierarchical structure of HPO. For each selected concept, a hierarchy recognition instance was constructed using a standardized prompt of the form:

> “*You are a medical expert. Please select the most accurate hypernym for the medical term ‘[query term]’.*”

Each instance consisted of four candidate parent concepts, exactly one of which corresponded to the true direct parent of the query concept in the HPO hierarchy. The construction of both correct answers and distractors followed a set of explicit design principles, described below.

1. Selection of true parent concepts. For each concept included in the evaluation set, the correct answer was defined as its immediate parent in the HPO is-a hierarchy. Restricting the task to direct parent relationships avoids ambiguity introduced by distant ancestors and ensures that each query has a single, well-defined correct answer.
2. Distractor design. Each instance included three distractors designed to challenge different aspects of hierarchical reasoning: (i) a sibling distractor, corresponding to a concept that shares the same parent as the query concept; (ii) a lexical distractor, lexically similar to the query term but occupying a different position in the ontology; and (iii) a random distractor, sampled from an unrelated branch of the ontology. Together, these distractors ensure that correct performance requires distinguishing true hierarchical relations from semantic proximity and surface-level similarity.
3. Task format. All instances were formatted as four-option multiple-choice questions using a fixed and standardized prompt template. The order of answer options was randomized for each instance. Models were instructed to return the option corresponding to the most appropriate parent concept.

### Task formulation for concept meaning understanding

The concept meaning understanding task was designed to evaluate whether large language models can correctly associate a medical term with its underlying semantic definition, rather than merely recognizing alternative names or hierarchical relations. In biomedical ontologies, concept meaning is formalized through natural language definitions that specify the essential characteristics of a concept; this task therefore probes definition-level concept understanding.

Building on the curated concept set described above, we formulated this task as a controlled multiple-choice definition selection problem grounded in the Human Phenotype Ontology. For each selected concept, a definition selection instance was constructed using a standardized prompt of the form:

> “*You are a medical expert. Please select the most accurate definition for the medical term ‘[query term]’.*”

The query term was deliberately chosen from among the non-preferred synonyms of the target concept, rather than its preferred name. In contrast, all candidate definition sentences were expressed using the concept’s preferred term. For example, a model might be queried with the term “Reactive airway disease” and asked to identify, from a set of candidate definitions, the correct definition corresponding to the concept Asthma as encoded in HPO. All candidate definitions followed a standardized format (“The definition of [the concept’s preferred term] is …”), with definition texts drawn verbatim from the ontology.

To construct a challenging and well-controlled set of candidate definitions under this formulation, we next describe how definition options were generated for each instance.

1. Candidate definition construction. Each instance consisted of exactly twenty candidate definition sentences, one of which corresponded to the correct ontological definition of the target concept. To ensure semantic plausibility among distractors, candidate definitions were constructed using a retrieval-based strategy. All HPO definition sentences were embedded using the Qwen3-Embedding-8B text embedding model^38^ and indexed in the Chroma vector database. For each target concept, its own definition was first excluded from the retrieval pool, and the nineteen most semantically similar definition sentences were retrieved based on cosine similarity. The correct definition of the target concept was then added to this set, resulting in a total of twenty candidate definitions.
2. Contextual knowledge conditions. To examine the influence of external ontological knowledge on definition-level understanding, this task was evaluated under three parallel context conditions. In the baseline context setting, no explicit ontological guidance was provided beyond the query and candidate definitions. In the augmented setting, models were supplied with accurate ontology-based information explicitly indicating that the query term and the preferred term appearing in the correct definition refer to the same concept. In the poisoned setting, models were instead provided with incorrect ontological information indicating that the query term and the preferred term do not refer to the same concept.
3. Task format. All instances were formatted as multiple-choice questions using a fixed and standardized prompt template. The order of candidate definition sentences was randomized for each instance. Models were instructed to return the index of the most accurate definition sentence.

### Large Language Model selection and inference settings

To evaluate medical concept understanding across contemporary LLMs while maintaining interpretability and experimental control, we selected a representative set of LLMs spanning commercial, open-source, and domain-specialized medical systems. This selection was guided by the goal of covering major model development paradigms—rather than exhaustively benchmarking all available models—and enabling principled comparisons across differences in accessibility, training regimes, and medical domain adaptation.

Specifically, we evaluated two commercial large language models, Gemini-3 Flash and GPT-5.2, released by Google and OpenAI, respectively. These models were selected as representative proprietary systems that are widely deployed, well-documented, and frequently used as reference points in prior studies of general-purpose and medical language understanding. Both models have demonstrated strong application-level performance across a range of medical and healthcare-oriented benchmarks, including HealthBench^39^, providing independent evidence that they operate near the current upper bound of real-world deployment capability. Importantly, our objective was not to compare all commercial models, but to include stable and reproducible proprietary baselines against which open-source and domain-specialized models could be meaningfully assessed.

In parallel, we included two state-of-the-art open-source general-purpose models, Qwen3-235B^40^ and DeepSeek-3.2^41^. These models represent high-capacity openly available LLMs that have been widely adopted in recent academic research and applied evaluations, including studies involving biomedical and scientific language tasks. Their inclusion enables assessment of whether conceptual understanding comparable to that of proprietary systems can be achieved under open training and deployment paradigms, without attempting to survey the full open-source model landscape.

To examine the effect of medical-domain adaptation, we further evaluated two domain-knowledge–tuned medical language models, Baichuan-M2^42^ and Citrus-Qwen72B^43^, both of which are fine-tuned on large-scale medical corpora and have reported competitive performance on medical evaluation benchmarks such as HealthBench. Crucially, these models share a common base architecture, Qwen2.5-72B, which was also included in all experiments. This controlled design allows observed performance differences to be attributed more directly to medical-domain fine-tuning, rather than to confounding factors such as architectural variation or model scale.

Across all tasks, models were evaluated using identical prompts and task formulations, under both standard inference settings and, where supported, reasoning-enabled settings (“thinking mode”) ^44^. All inference was conducted with default or recommended decoding parameters provided by model developers, and no task-specific prompt engineering or post hoc optimization was applied. This uniform evaluation protocol ensures that observed differences in performance reflect intrinsic differences in model behavior and conceptual representations, rather than artifacts of prompt design or inference configuration.

### Evaluation metrics

Across all tasks, model performance was evaluated using accuracy, defined as the proportion of instances in which the model selected the correct answer. Because all tasks were formulated as fixed-option multiple-choice questions with exactly one correct option per instance, accuracy provides a direct, interpretable, and task-consistent measure of performance. Accuracy was computed as:

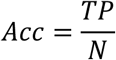

where *N* denotes the total number of evaluation instances and *TP* denotes the number of instances correctly answered by the model.

For the concept identity and concept hierarchy tasks, each instance consisted of four candidate options, yielding a chance-level accuracy of 25%. For the concept meaning task, each instance consisted of twenty candidate definition sentences, corresponding to a chance-level accuracy of 5%. Accuracy was computed independently for each model, task, and experimental condition (e.g., with or without thinking mode; baseline, augmented, or poisoned context).

Given the different chance baselines and task formulations, we report task-specific accuracy values throughout and do not aggregate results across tasks. All reported metrics therefore reflect task-specific performance rather than normalized or composite scores.

### Statistical analysis

Statistical significance was assessed using McNemar’s test on paired predictions^45^, comparing model performance on the same set of test instances. This test is appropriate for evaluating differences between paired classifiers on binary outcomes (correct versus incorrect) without assuming underlying score distributions.

McNemar’s test was applied to pairwise comparisons within each task, including comparisons between standard and reasoning-enabled settings for the same model, as well as selected model-to-model comparisons under identical experimental conditions. To account for multiple comparisons, Bonferroni correction^46^ was applied within each task.

All statistical tests were two-sided, and differences were considered statistically significant at a corrected p-value threshold of 0.05.

## Data Availability

All data produced in the present study are available upon reasonable request to the authors

## Acknowledgments

We sincerely thank colleagues from the Excellent Innovation Team of the Philosophy and Social Sciences in the Universities and Colleges of Jiangsu Province “The Public Health Policy and Management Innovation Research Team” for their thoughtful suggestions to improve this work.

## Author contributions

**L.D**: Writing – review & editing, Writing – original draft, Supervision, Project administration, Methodology, Funding acquisition, Conceptualization. **L.C**: Writing – review & editing, Visualization, Validation, Resources, Methodology, Formal analysis, Data curation, Conceptualization. **M.L**: Writing – review & editing, Validation, Investigation, Data curation.

